# Previous and active tuberculosis increases risk of death and prolongs recovery in patients with COVID-19

**DOI:** 10.1101/2020.07.22.20154575

**Authors:** Karla Therese L. Sy, Nel Jason L. Haw, Jhanna Uy

**Affiliations:** Department of Epidemiology, Boston University School of Public Health, Boston MA, United States; Department of Global Health, Boston University School of Public Health, Boston MA, United States; Health Sciences Program, School of Science and Engineering, Ateneo de Manila University, Manila, Philippines; Philippine Institute for Development Studies, Manila, Philippines

**Keywords:** COVID-19, SARS-CoV-2, Tuberculosis, Philippines

## Abstract

**Background:** There is a growing literature on the association of SARS-CoV-2 and other chronic conditions, such as noncommunicable diseases. However, little is known about the impact of coinfection with tuberculosis. We aimed to compare the risk of death and recovery, as well as time-to-death and time-to-recovery, in COVID-19 patients with and without TB.

**Methods:** We created a 4:1 propensity score matched sample of COVID-19 patients without and with tuberculosis, using COVID-19 surveillance data in the Philippines. We conducted a longitudinal cohort analysis of matched COVID-19 patients as of May 17, 2020, following them until June 15, 2020. The primary analysis estimated the risk ratios of death and recovery in patients with and without tuberculosis. Kaplan-Meier curves described time-to-death and time-to-recovery stratified by tuberculosis status, and differences in survival were assessed using the Wilcoxon test.

**Results:** The risk of death in COVID-19 patients with tuberculosis was 2.17 times higher than in those without (95% CI: 1.40-3.37). The risk of recovery in COVID-19 patients with tuberculosis was 25% lower than in those without (RR=0.75, 95% CI 0.63-0.91). Similarly, time-to-death was significantly shorter (p=0.0031) and time-to-recovery significantly longer in patients with tuberculosis (p=0.0046).

**Conclusions:** Our findings show that coinfection with tuberculosis increased morbidity and mortality in COVID-19 patients. Our findings highlight the need to prioritize routine and testing services for tuberculosis, although health systems are disrupted by the heavy burden of the SARS-CoV-2 pandemic.

## Introduction

As of July 2020, the global burden of the COVID-19 pandemic has reached 15 million cases, and has caused large-scale outbreaks in many countries [1]. The COVID-19 pandemic is a substantial strain to healthcare systems worldwide, particularly in resource-limited settings with high prevalence of comorbid conditions such as tuberculosis (TB). TB is a major cause of morbidity and mortality, and caused 1.4 million deaths in 2018, the main global infectious cause of death [2]. TB disproportionately affects low-and-middle income countries, where TB epidemics are fueled by coinfection with HIV/AIDS [3] and multidrug resistance [4, 5]. The pandemic has caused disruptions of TB treatment and programs [6, 7], which have disproportionately impacted socially disadvantaged communities with TB [8]. In particular, fewer patients have sought medical treatment for TB, research on TB has been reduced, availability of drugs against TB has decreased, and there are issues regarding food supply and nutritional support during the pandemic [9].

The COVID-19 outbreak in the Philippines is a continuing public health crisis, with 12,513 reported cases as of May 17, 2020. TB is also a major public health problem in the Philippines with the third highest incidence of TB globally, and one million individuals have active TB [10]. Most of the research on SARS-CoV-2 has been conducted in high-income countries, and studies suggest worse outcomes in COVID-19 patients with other respiratory diseases, such as chronic obstructive pulmonary disease (COPD) [11, 12] and asthma [13]. However, little is known about the impact of coinfection with TB on COVID-19 outcomes. Pulmonary TB causes destruction of lung parenchyma [14], which may predispose COVID-19 patients with TB to aggressive disease and high mortality. We hypothesized that COVID-19 patients with previous or active TB may have worse clinical outcomes than those without. This study compares the risk of and time-to-death and recovery in COVID-19 patients with and without TB coinfection in the Philippines.

## Materials and Methods

### Data

Data was obtained from the Philippine national COVID-19 surveillance implemented and managed by the Philippine Department of Health Epidemiology Bureau (DOH-EB). All data on COVID-19 cases was de-identified, and data came from continuing surveillance efforts by DOH-EB as part of an ongoing outbreak investigation. Thus, institutional review board approval was deemed unnecessary. Part of the case data is publicly available through the DOH DataDrop initiative [15].

### Study Population

We included all reported COVID-19 cases in the Philippines as of May 17, 2020, following them until June 15, 2020. COVID-19 cases were confirmed with positive real-time reverse transcription polymerase chain reaction (RT-PCR) from laboratories accredited by the DOH and the Research Institute for Tropical Medicine (RITM) [16]. Information on confirmed cases were collected using structured interviews by the physician or nurse on duty at initial consultation as part of routine surveillance using a case investigation form (CIF), filled out manually or electronically. We used propensity score matching to create similar populations of COVID-19 patients with and without previous and active TB. To create the propensity scores, we predicted the risk of having TB using logistic regression with the potential confounders age, sex, and other comorbid conditions (COPD, asthma, diabetes, hypertension, cancer, renal disease, cardiac disease, and autoimmune disorders). We matched one COVID-19 patient with TB to four without, using nearest neighbor matching of propensity scores, a caliper of 0.05, and with no replacement. For a secondary analysis, we created a propensity score matched subsample of COVID-19 patients admitted to hospital.

### Exposure

Our main exposure of interest was confirmed TB, which was defined as a history of or a current diagnosis of TB. Comorbidity data and death certificate data encoded as part of the CIF were used to determine the status of the TB infection.

### Outcomes

The two main outcomes of interest were (1) death and (2) recovery. We modelled this as a dichotomous (yes/no) variable, as well as a time-to-event variable. The time-to-event variables for both primary outcomes were the time from symptom onset to death or recovery. Death was defined as deaths during active COVID-19 and declared as a death by the DOH-EB. Recovery was defined as cases declared recovered by the DOH-EB based on criteria for testing, clinical improvement, and additional days of quarantine. We assessed the time from hospital admission to discharge in patients admitted to hospital with dates validated in medical records. Another secondary outcome was the relative risk of admission in the propensity score matched sample.

### Statistical analyses

Descriptive statistics were used to characterize the sample. Student’s t-test was used to assess differences in age between patients with and without TB. Pearson chi-square test was used to examine differences in gender, comorbidities, health status and admission status. The primary analysis estimated the relative risks (risk ratios) of death and recovery with a modified Poisson regression with a robust variance estimator and a log link.

Kaplan-Meier curves stratified by TB status were used to plot survival curves of the time-to-event variables. Non-parametric analysis of differences in survival between patients with and without TB was done using the Wilcoxon test. Individuals were censored at their last date of follow-up if they did not have the outcome of interest. For the analysis of time-to-recovery, we censored subjects who were dead on the last day of administrative follow-up for the whole sample. This was done to not raise the bias estimate of recovery by removing patients who died as censored observations. All statistical analyses were conducted in R 4.0.0 [17].

## Results

### Propensity score matching

The initial unmatched sample consisted of 12,513 COVID-19 patients, of which 113 (1.0%) had confirmed TB. Sex differed significantly between patients with and without TB (p=0.001), as well as hypertension (p<0.001) and diabetes (p<0.006) (**Supplementary Table 1**). We removed all patients with missing covariates on variables from the propensity score matching, including seven TB patients, and 4,510 COVID-19 patients were included in the final propensity score-matched sample. The final matched sample consisted of 530 patients, with 106 cases with TB cases and 424 without. Matching successfully created populations with similar baseline characteristics (all p>0.05; **Table 1**). The mean age of the total sample was 48.9 years (SD=18.88; 95% CI: 47.2-50.5), 68.9% were males, 13.4% (n=71) died, and 67.7% (n=359) recovered. The unmatched sample of COVID-19 patients admitted to hospital consisted of 3,869 people, of which 70 (1.8%) had confirmed TB. In the propensity score matched cohort of admitted patients, three TB patients had missing covariates, and one TB patient had no control match. The final subsample included 66 TB patients and 264 matched controls with similar baseline characteristics (all p>0.05; **Table 1**).

**Table 1.** Demographics and health status of the matched full sample and matched admitted sample by TB status.

| **Matched sample** | **Overall (n=530)** | **No TB (n=424)** | **TB (n=106)** | **P** |
| --- | --- | --- | --- | --- |
| **Age (mean (SD))** | 48.86 (18.88) | 48.84 (18.71) | 48.92 (19.63) | 0.968 |
| **Male (%)** | 373 (70.4) | 300 (70.8) | 73 (68.9) | 0.794 |
| **Comorbidities (%)** |  |  |  |  |
| **Hypertension** | 109 (20.6) | 87 (20.5) | 22 (20.8) | 1.000 |
| **Diabetes** | 67 (12.6) | 53 (12.5) | 14 (13.2) | 0.974 |
| **Cancer** | 6 (1.1) | 5 (1.2) | 1 (0.9) | 1.000 |
| **Renal Cancer** | 16 (3.0) | 12 (2.8) | 4 (3.8) | 0.849 |
| **Cardiac disease** | 41 (7.7) | 33 (7.8) | 8 (7.5) | 1.000 |
| **Asthma** | 21 (4.0) | 17 (4.0) | 4 (3.8) | 1.000 |
| **COPD** | 6 (1.1) | 3 (0.7) | 3 (2.8) | 0.182 |
| **Autoimmune disease** | 0 (0) | 0 (0) | 0 (0) | 1.000 |
| **Health Status (%)** |  |  |  |  |
| **Died** | 71 (13.4) | 46 (10.8) | 25 (23.6) | 0.001 |
| **Recovered** | 359 (67.7) | 302 (71.2) | 57 (53.8) | 0.001 |
| **Admission Status (%)** |  |  |  | 0.038 |
| **Admitted** | 303 (57.2) | 236 (55.7) | 67 (63.2) |  |
| **Not Admitted** | 162 (30.6) | 140 (33.0) | 22 (20.8) |  |
| **Unknown** | 65 (12.3) | 48 (11.3) | 17 (16.0) |  |
| **Admitted sample** | **Overall (n=330)** | **No TB (n=264)** | **TB (n=66)** | **P** |
| **Age (mean (SD))** | 49.50 (18.54) | 49.29 (17.71) | 50.33 (21.68) | 0.683 |
| **Male (%)** | 235 (71.2) | 187 (70.8) | 48 (72.7) | 0.879 |
| **Comorbidities (%)** |  |  |  |  |
| **Hypertension** | 80 (24.2) | 64 (24.2) | 16 (24.2) | 1.000 |
| **Diabetes** | 51 (15.5) | 41 (15.5) | 10 (15.2) | 1.000 |
| **Cancer** | 3 (0.9) | 2 (0.8) | 1 (1.5) | 1.000 |
| **Renal Cancer** | 13 (3.9) | 9 (3.4) | 4 (6.1) | 0.524 |
| **Cardiac disease** | 27 (8.2) | 20 (7.6) | 7 (10.6) | 0.581 |
| **Asthma** | 1 (0.3) | 0 (0.0) | 1 (1.5) | 0.453 |
| **COPD** | 1 (0.3) | 0 (0.0) | 1 (1.5) | 0.453 |
| **Autoimmune disease** | 0 (0) | 0 (0) | 0 (0) | 1.000 |
| **Health Status (%)** |  |  |  |  |
| **Died** | 50 (15.2) | 32 (12.1) | 18 (27.3) | 0.004 |
| **Recovered** | 218 (66.1) | 180 (68.2) | 38 (57.6) | 0.138 |

Patients with TB had 2.17 times higher risk of death than those without (95% CI: 1.40-3.37). When assessed in hospitalized patients, those with TB had a similar higher risk of death (RR=2.25, 95% CI: 1.35-3.75). The risk of recovery in patients with TB was 25% lower than in those without (RR=0.75, 0.63-0.91). The risk of recovery was similar in admitted patients, but not significant (RR=0.84, 95% CI: 0.68-1.06). There was a 20% higher risk of admission in patients with TB (RR=1.20, 95% CI: 1.04-1.38) (**Table 2**).

**Table 2.** Relative risks of death, recovery and admission in full matched sample and matched admitted sample.

|  | **RR (95% CI)** | ***P*** |
| --- | --- | --- |
| **Matched cohort (n=530)** |  |  |
| **Death** | 2.17 (1.4-3.37) | 0.001 |
| **Recovery** | 0.75 (0.63-0.91) | 0.003 |
| **Admission** | 1.20 (1.04-1.38) | 0.012 |
| **Admitted only (n=330)** |  |  |
| **Death** | 2.25 (1.35-3.75) | 0.002 |
| **Recovery** | 0.84 (0.68-1.06) | 0.137 |

Kaplan-Meier survival analysis (Figure 1) showed that the time-to-death in patients with TB was significantly shorter than in those without for both the propensity score matched sample (p=0.0031) and in the subsample of admitted patients (p=0.0052). The time-to-recovery in patients with TB was significantly longer than in those without (full sample p=0.0046; admitted only p=0.02) (**Figure 1**). The time-to-admission did not differ significantly (p=0.17).

**Figure 1.**
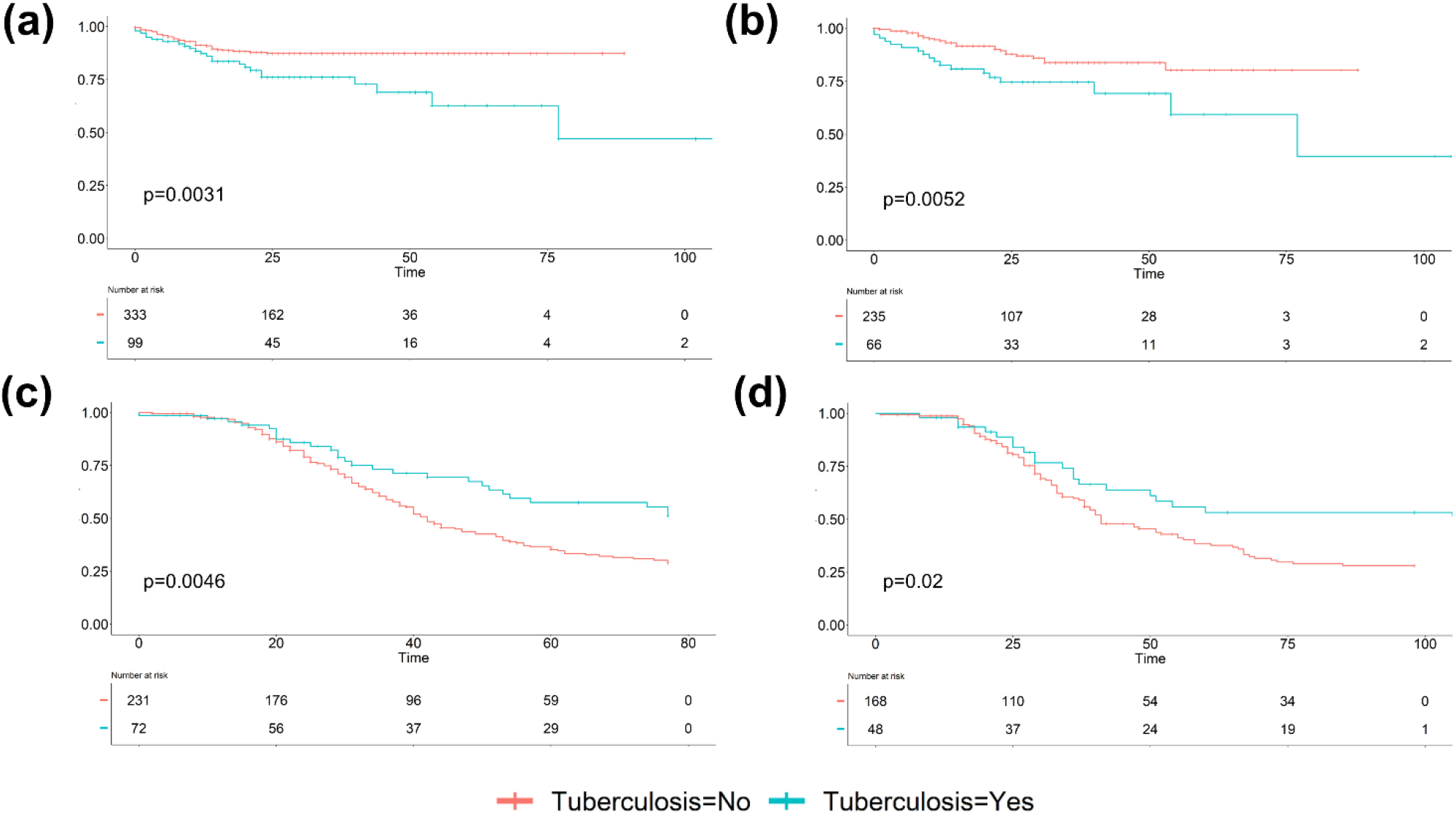
Survival analysis for (a) time-to-death in whole matched sample (b) time-to-death in hospitalized subsample (c) time-to-recovery in whole matched sample (d) time-to-recovery in hospitalized subsample.

## Discussion

The ongoing SARS-CoV-2 pandemic poses a challenge for TB prevention and treatment worldwide. Our findings showed that COVID-19 patients with TB had a two-fold increased risk of death, and were less likely to recover. Moreover, time-to-death was shorter and time-to-recovery longer than in patients without TB. Given that SARS-CoV-2 has caused a substantial number of deaths worldwide (greater than 600,000 deaths by July 2020) [1], some of the deaths can be attributable to the increased risk of death and slower recovery in COVID-19 patients with TB. These deaths can be prevented by continuing to provide essential TB services, particularly in countries with high TB burden, where TB cases need to continue to be correctly diagnosed and promptly treated. Moreover, our finding suggests that areas with greater TB burden would have higher case fatality rates of COVID-19, which is important for public health planning and resource allocation. As COVID-19 cases are continuing to increase in many low and middle income countries with high TB incidence, such as countries in Southern Africa, Latin America, and Asia [1], mitigation strategies need to target individuals with TB in order to reduce the global burden of COVID-19.

Our findings are consistent with preliminary findings from the Western Cape province of South Africa, where COVID-19 patients with a current diagnosis had a 2.5 times higher risk of death, and those with previous TB had a 50% higher risk [18]. Other studies have examined the association between TB and COVID-19; a case-control study in China demonstrated an increased susceptibility of TB patients to SARS-CoV-2 infection, and increased severity of symptom development [19]. Several studies have also described the interrelationship of these two diseases, such as assessing deaths in patients with TB and COVID-19 co-infection [20] and the post-TB sequalae in a global cohort of TB patients with COVID-19 [21]. However, studies with longer follow-ups and adequate sample size needed to make inferential comparisons are limited. To our knowledge, this is the first longitudinal cohort study investigating the association between COVID-19 and TB with sufficient sample size in a high-burden TB setting.

There are a number of limitations of the study. We used propensity score matching to adjust for confounding factors. However, residual confounding could still be an issue for variables we did not match. Specifically, we had no data on HIV/AIDs in our cohort, and this might have biased our estimates since patients with TB are more likely to be infected with HIV. However, propensity score matching on all specified comorbid conditions would reduce confounding substantially, and any potential residual confounding would be limited. Moreover, as this data was obtained from the Philippine national surveillance of COVID-19, some dates for the time-to-event outcomes were missing. In survival analyses, we excluded individuals with missing dates, and conducted statistical tests to ensure that the confounder distribution of the included sample for each time-to-event analysis did not differ significantly between patients with and without TB (**Supplementary Table 2**). In addition, the main outcomes death and recovery could potentially have some missing data; the survival analysis accounted for missing data by censoring on the last day of follow-up for patients without the outcome. Furthermore, since the data was collected as part of a larger surveillance effort, previous TB diagnosis and current TB disease were aggregated as confirmed TB, and we were unable to distinguish between the effects of these two groups; even so, the consistently strong effect size demonstrates the consequence of TB and COVID-19 co-infection, adding to the body of evidence that TB worsens the prognosis of COVID-19 patients. Additionally, in the interest of rapid reporting of scientific findings during a pandemic, the patients were only followed up for a short period of time, and we were only able to determine the short-term outcomes in TB cases with COVID-19. Future longitudinal cohort studies need to establish the long-term outcomes among those coinfected with TB and COVID-19.

Our results demonstrated a direct relationship between COVID-19 and TB, but SARS-CoV-2 also indirectly contributes to TB-related morbidity and mortality. COVID-19 has and will continue to cause a substantial strain to healthcare systems worldwide, and high mortality is predicted for TB patients due to TB program disruptions, diagnostic delays, treatment interruptions, and lack of access to drugs [6, 7, 9]. Moreover, inaccessibility of health facilities and reduced ability to pay for medical costs disproportionately affect socially disadvantaged populations, compounding already existing health inequalities, particularly in countries with already vulnerable health and economic systems [22]. Our findings highlight the need to continue prioritizing routine and testing services for TB, despite disruptions in health and social systems during the COVID-19 pandemic. Moreover, since a large proportion of patients with TB are coinfected with HIV, further research on the co-occurrence of these three infections, as well as on other endemic communicable and noncommunicable diseases in high TB-burden countries, is important for accurate assessment of the global burden of SARS-CoV-2. Future studies with focus on the relationship between TB and COVID-19 are warranted to support appropriate planning and resource allocation, as SARS-CoV-2 continues to spread around the world.

## Data Availability

Data are publicly available through through the Philippine Department of Health DataDrop initiative (https://www.doh.gov.ph/covid19tracker).

## Author contributions

K. Sy contributed to conceptualization. K. Sy, N. Haw, and J. Uy contributed to data acquisition. K. Sy contributed to data analysis. All authors contributed to interpretation of results and manuscript writing.

## Supplementary Material

**Supplementary Table 1.** Demographics and health status of the unmatched full sample and unmatched admitted sample.

| Full sample | Overall<br>(n=12,513) | No TB (n=4407) | TB (n=113) | *P* |
| --- | --- | --- | --- | --- |
| **Age (mean (SD))** | 44.21 (18.18) | 49.64 (17.76) | 49.15 (19.62) | 0.774 |
| **Male (%)** | 6792 (54.3) | 2346 (53.2) | 80 (70.8) | <0.001 |
| **Comorbidities (%)** |  |  |  |  |
| Hypertension | 1601 (35.5) | 1579 (35.8) | 22 (20.8) | 0.002 |
| Diabetes | 1137 (25.2) | 1123 (25.5) | 14 (13.2) | 0.006 |
| Cancer | 89 (2.0) | 88 (2.0) | 1 (0.9) | 0.676 |
| Renal Cancer | 219 (4.9) | 215 (4.9) | 4 (3.8) | 0.768 |
| Cardiac disease | 285 (6.3) | 277 (6.3) | 8 (7.5) | 0.745 |
| Asthma | 292 (6.5) | 288 (6.5) | 4 (3.8) | 0.346 |
| COPD | 37 (0.8) | 34 (0.8) | 3 (2.8) | 0.075 |
| Autoimmune disease | 9 (0.2) | 9 (0.2) | 0 (0.0) | 1.000 |
| **Health Status (%)** |  |  |  |  |
| Died | 980 (7.8) | 546 (12.4) | 32 (28.3) | <0.001 |
| Recovered | 5867 (46.9) | 3110 (70.6) | 57 (50.4) | <0.001 |
| **Admission Status (%)** |  |  |  | 0.001 |
| Admitted | 3869 (30.9) | 2394 (54.3) | 70 (61.9) |  |
| Not Admitted | 2656 (21.2) | 1540 (34.9) | 22 (19.5) |  |
| Unknown | 5988 (47.9) | 473 (10.7) | 21 (18.6) |  |
| Admitted sample | Overall (n=3869) | No TB (n=2394) | TB (n=70) | *P* |
| **Age (mean (SD))** | 50.47 (18.30) | 52.75 (17.90) | 50.37 (21.17) | 0.275 |
| **Male (%)** | 2168 (56.0) | 1344 (56.2) | 51 (72.9) | 0.008 |
| **Comorbidities (%)** |  |  |  |  |
| Hypertension | 977 (39.7) | 961 (40.1) | 16 (23.9) | 0.011 |
| Diabetes | 749 (30.4) | 739 (30.9) | 10 (14.9) | 0.008 |
| Cancer | 67 (2.7) | 66 (2.8) | 1 (1.5) | 0.805 |
| Renal Cancer | 159 (6.5) | 155 (6.5) | 4 (6.0) | 1.000 |
| Cardiac disease | 210 (8.5) | 203 (8.5) | 7 (10.4) | 0.729 |
| Asthma | 164 (6.7) | 163 (6.8) | 1 (1.5) | 0.141 |
| COPD | 30 (1.2) | 28 (1.2) | 2 (3.0) | 0.441 |
| Autoimmune disease | 6 (0.2) | 6 (0.3) | 0 (0.0) | 1.000 |
| **Health Status (%)** |  |  |  |  |
| Died | 601 (15.5) | 401 (16.8) | 22 (31.4) | 0.002 |
| Recovered | 2639 (68.2) | 1650 (69.0) | 38 (54.3) | 0.013 |

**Supplementary Table 2.** Confounder distribution of each time-to-event analysis.

| **Time-to-death** |  |  |  |  |
| --- | --- | --- | --- | --- |
| **Matched sample** | **Overall**<br>**(n=432)** | **No TB (n=333)** | **TB (n=99)** | **P** |
| **Age (mean (SD))** | 49.52 (18.82) | 49.83 (18.53) | 48.51 (19.82) | 0.540 |
| **Male (%)** | 304 (70.4) | 236 (70.9) | 68 (68.7) | 0.770 |
| **Comorbidities (%)** |  |  |  |  |
| **Hypertension** | 93 (21.5) | 73 (21.9) | 20 (20.2) | 0.821 |
| **Diabetes** | 59 (13.7) | 46 (13.8) | 13 (13.1) | 0.994 |
| **Cancer** | 4 (0.9) | 3 (0.9) | 1 (1.0) | 1.000 |
| **Renal Cancer** | 13 (3.0) | 9 (2.7) | 4 (4.0) | 0.727 |
| **Cardiac disease** | 37 (8.6) | 29 (8.7) | 8 (8.1) | 1.000 |
| **Asthma** | 21 (4.9) | 17 (5.1) | 4 (4.0) | 0.868 |
| **COPD** | 6 (1.4) | 3 (0.9) | 3 (3.0) | 0.271 |
| **Autoimmune disease** | 432 (100.0) | 333 (100.0) | 99 (100.0) | 1.000 |
| **Admitted sample** | **Overall**<br>**(n=301)** | **No TB (n=235)** | **TB (n=66)** | **P** |
| **Age (mean (SD))** | 49.79 (18.53) | 49.64 (17.59) | 50.33 (21.68) | 0.789 |
| **Male (%)** | 218 (72.4) | 170 (72.3) | 48 (72.7) | 1.000 |
| **Comorbidities (%)** |  |  |  |  |
| **Hypertension** | 75 (24.9) | 59 (25.1) | 16 (24.2) | 1.000 |
| **Diabetes** | 48 (15.9) | 38 (16.2) | 10 (15.2) | 0.992 |
| **Cancer** | 3 (1.0) | 2 (0.9) | 1 (1.5) | 1.000 |
| **Renal Cancer** | 12 (4.0) | 8 (3.4) | 4 (6.1) | 0.536 |
| **Cardiac disease** | 24 (8.0) | 17 (7.2) | 7 (10.6) | 0.524 |
| **Asthma** | 1 (0.3) | 0 (0.0) | 1 (1.5) | 0.497 |
| **COPD** | 1 (0.3) | 0 (0.0) | 1 (1.5) | 0.497 |
| **Autoimmune disease** | 301 (100.0) | 235 (100.0) | 66 (100.0) | 1.000 |
| **Time-to-recovery** |  |  |  |  |
| **Matched sample** | **Overall**<br>**(n=303)** | **No TB (n=231)** | **TB (n=72)** | **P** |
| **Age (mean (SD))** | 50.49 (19.41) | 51.07 (18.84) | 48.62 (21.16) | 0.352 |
| **Male (%)** | 215 (71.0) | 167 (72.3) | 48 (66.7) | 0.441 |
| **Comorbidities (%)** |  |  |  |  |
| **Hypertension** | 62 (20.5) | 48 (20.8) | 14 (19.4) | 0.938 |
| **Diabetes** | 44 (14.5) | 35 (15.2) | 9 (12.5) | 0.714 |
| **Cancer** | 3 (1.0) | 2 (0.9) | 1 (1.4) | 1.000 |
| **Renal Cancer** | 12 (4.0) | 8 (3.5) | 4 (5.6) | 0.654 |
| **Cardiac disease** | 33 (10.9) | 25 (10.8) | 8 (11.1) | 1.000 |
| **Asthma** | 13 (4.3) | 10 (4.3) | 3 (4.2) | 1.000 |
| **COPD** | 5 (1.7) | 3 (1.3) | 2 (2.8) | 0.741 |
| **Autoimmune disease** | 303 (100.0) | 231 (100.0) | 72 (100.0) | 1.000 |
| **Time-to-recovery** |  |  |  |  |
| **Admitted sample** | **Overall**<br>**(n=216)** | **No TB (n=168)** | **TB (n=48)** | **P** |
| **Age (mean (SD))** | 50.02 (19.25) | 50.24 (18.01) | 49.27 (23.27) | 0.760 |
| **Male (%)** | 158 (73.1) | 123 (73.2) | 35 (72.9) | 1.000 |
| **Comorbidities (%)** |  |  |  |  |
| **Hypertension** | 59 (27.3) | 49 (29.2) | 10 (20.8) | 0.338 |
| **Diabetes** | 37 (17.1) | 31 (18.5) | 6 (12.5) | 0.454 |
| **Cancer** | 2 (0.9) | 1 (0.6) | 1 (2.1) | 0.924 |
| **Renal Cancer** | 9 (4.2) | 5 (3.0) | 4 (8.3) | 0.219 |
| **Cardiac disease** | 24 (11.1) | 17 (10.1) | 7 (14.6) | 0.543 |
| **Asthma** | 1 (0.5) | 0 (0.0) | 1 (2.1) | 0.503 |
| **COPD** | 1 (0.5) | 0 (0.0) | 1 (2.1) | 0.503 |
| **Autoimmune disease** | 216 (100.0) | 168 (100.0) | 48 (100.0) | 1.000 |

**Time-to-admission**
| **Matched sample** | **Overall**<br>**(n=441)** | **No TB (n=341)** | **TB (n=100)** | ***P*** |
| --- | --- | --- | --- | --- |
| **Age (mean (SD))** | 49.90 (18.87) | 50.26 (18.60) | 48.68 (19.80) | 0.463 |
| **Male (%)** | 311 (70.5) | 243 (71.3) | 68 (68.0) | 0.614 |
| **Comorbidities (%)** |  |  |  |  |
| **Hypertension** | 94 (21.3) | 74 (21.7) | 20 (20.0) | 0.821 |
| **Diabetes** | 61 (13.8) | 48 (14.1) | 13 (13.0) | 0.913 |
| **Cancer** | 4 (0.9) | 3 (0.9) | 1 (1.0) | 1.000 |
| **Renal Cancer** | 14 (3.2) | 10 (2.9) | 4 (4.0) | 0.833 |
| **Cardiac disease** | 40 (9.1) | 32 (9.4) | 8 (8.0) | 0.821 |
| **Asthma** | 21 (4.8) | 17 (5.0) | 4 (4.0) | 0.889 |
| **COPD** | 6 (1.4) | 3 (0.9) | 3 (3.0) | 0.263 |
| **Autoimmune disease** | 441 (100.0) | 341 (100.0) | 100 (100.0) | 1.000 |

